# Predictive power of wastewater for nowcasting infectious disease transmission: a retrospective case study of five sewershed areas in Louisville, Kentucky

**DOI:** 10.1101/2023.05.31.23290619

**Authors:** Fayette Klaassen, Rochelle H. Holm, Ted Smith, Ted Cohen, Aruni Bhatnagar, Nicolas A. Menzies

## Abstract

**Background:** Epidemiological nowcasting traditionally relies on count surveillance data. The availability and quality of such data may vary over time, limiting their representation of true infections. Wastewater data correlates with traditional surveillance data and may provide additional value for nowcasting disease trends.

**Methods:** We obtained SARS-CoV-2 case, death, wastewater, and serosurvey data for Jefferson County, Kentucky, between August 2020 and March 2021, and parameterized an existing nowcasting model using combinations of these data. We assessed the predictive performance and variability at the sewershed level and compared the effects of adding or replacing wastewater data to case and death reports.

**Findings:** Adding wastewater data minimally improved the predictive performance of nowcasts compared to a model fitted to case and death data (Weighted Interval Score (WIS) 0·208 versus 0·223), and reduced the predictive performance compared to a model fitted to deaths data (WIS 0·517 versus 0·500). Adding wastewater data to deaths data improved the nowcasts agreement to estimates from models using cases and deaths data. These findings were consistent across individual sewersheds as well as for models fit to the aggregated data of all 5 sewersheds. Retrospective reconstructions of epidemiological dynamics created using different combinations of data were in general agreement (coverage > 75%).

**Interpretation:** These findings show that wastewater data may be valuable for infectious disease nowcasting when clinical surveillance data are absent, such as early in a pandemic or in low-resource settings where systematic collection of epidemiologic data is difficult.

**Funding:** CDC, Louisville-Jefferson County Metro Government, and other funders.

## Introduction

Epidemiological nowcasting is an important tool for understanding infectious disease trends, and could be used to produce real-time estimates of the transmission rate and reproduction number (*Rt*) of an infectious pathogen.^1^ This information is critical for informing risk assessment and the subsequent public health response.

To-date, nowcasting methods rely on surveillance data (e.g., case reports, hospitalizations, vaccination records) that have limitations for describing disease trends.^2^ First, asymptomatic and mild infections may not be diagnosed, and so will be missing from clinical surveillance data. Second, variation in healthcare access or sampling methods can mean that available data are not representative of the population of interest.^3, 4^ A third limitation is the lag between the time transmission occurs and the time when the consequences of transmission become apparent in measured quantities. For instance, a COVID-19 diagnosis occurs only after symptoms have developed, around five days after infection with severe acute respiratory syndrome coronavirus 2 (SARS-COV-2).^5^ There may be additional lags between the development of symptoms and the time of diagnosis, and between diagnosis and reporting. Finally, surveillance data are often aggregated over large geographical areas or time periods, which may mask heterogeneities in transmission patterns and prevent prompt geographic targeting of mitigation measures. For example, in the initial stages of the COVID-19 pandemic, states and counties generally reported daily surveillance data, often by city, county and state. Over time reporting frequencies decreased to weekly or biweekly, reducing the temporal resolution of the data and increasing the lag between infection and reporting.

Wastewater-based epidemiology (WBE) has been used for public health monitoring of infectious diseases such as polio^6^ and was increasingly relied upon during the COVID-19 pandemic.^7^ Samples from wastewater provide passively collected information from the entire population attached to the sewer network, and if collected at a regular frequency, wastewater data can have a strong temporal correlation with changes in disease incidence.^8^ The promise of improved temporal signal in the wastewater data is a major potential benefit given the importance of accurate information on current transmission dynamics in informing prompt action or policy responses. In the United States (USA), several large, publicly accessible wastewater databases have been developed during the SARS-CoV-2 pandemic, as well as for tracking of other infectious diseases.^9–11^

Wastewater data has been used in nowcasts of large metropolitan areas, and it has been found that there is no definitive agreement on a case count to water concentration.^8, 12–14^ Nevertheless, WBE for surveillance has its own unique challenges, including variation in sampling and quantification methods, and challenges mapping wastewater levels to estimates of local disease burden.^15–17^ As a result, the value of wastewater data for nowcasting disease transmission in sub-populations in addition to, or instead of, traditional surveillance data remains unclear. To assess the added utility of using WBE to count-based nowcasting, we examine SARS-CoV-2 surveillance data from Louisville, KY, which were available for five distinct sewersheds and could be combined for an aggregate countywide view. We used these data and a published nowcasting model to evaluate the utility of wastewater data for improving the accuracy of nowcasts.

## Methods

### Data

We used data from Louisville/Jefferson County, Kentucky, covering the period between August 2020 and March 2021, prior to public vaccine access.^8^ Data were geocoded to five wastewater treatment plant catchment areas or “sewersheds”, named MSD0[1-5], that cover 97% of the county population (Figure 1 and Table S1). Data were aggregated for a countywide level for comparison.

**Figure 1.**
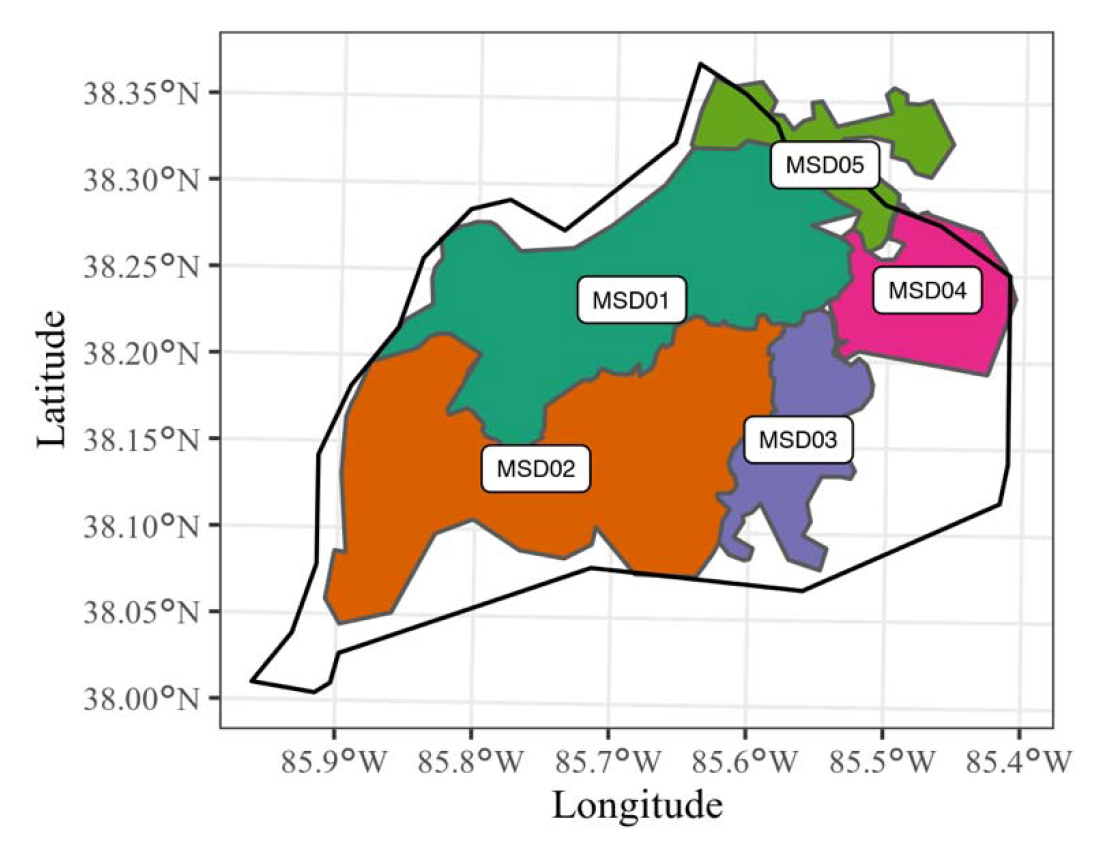
**Five studied wastewater treatment plant zones (sewersheds), Jefferson County, KY (USA).**

**Figure 2.**
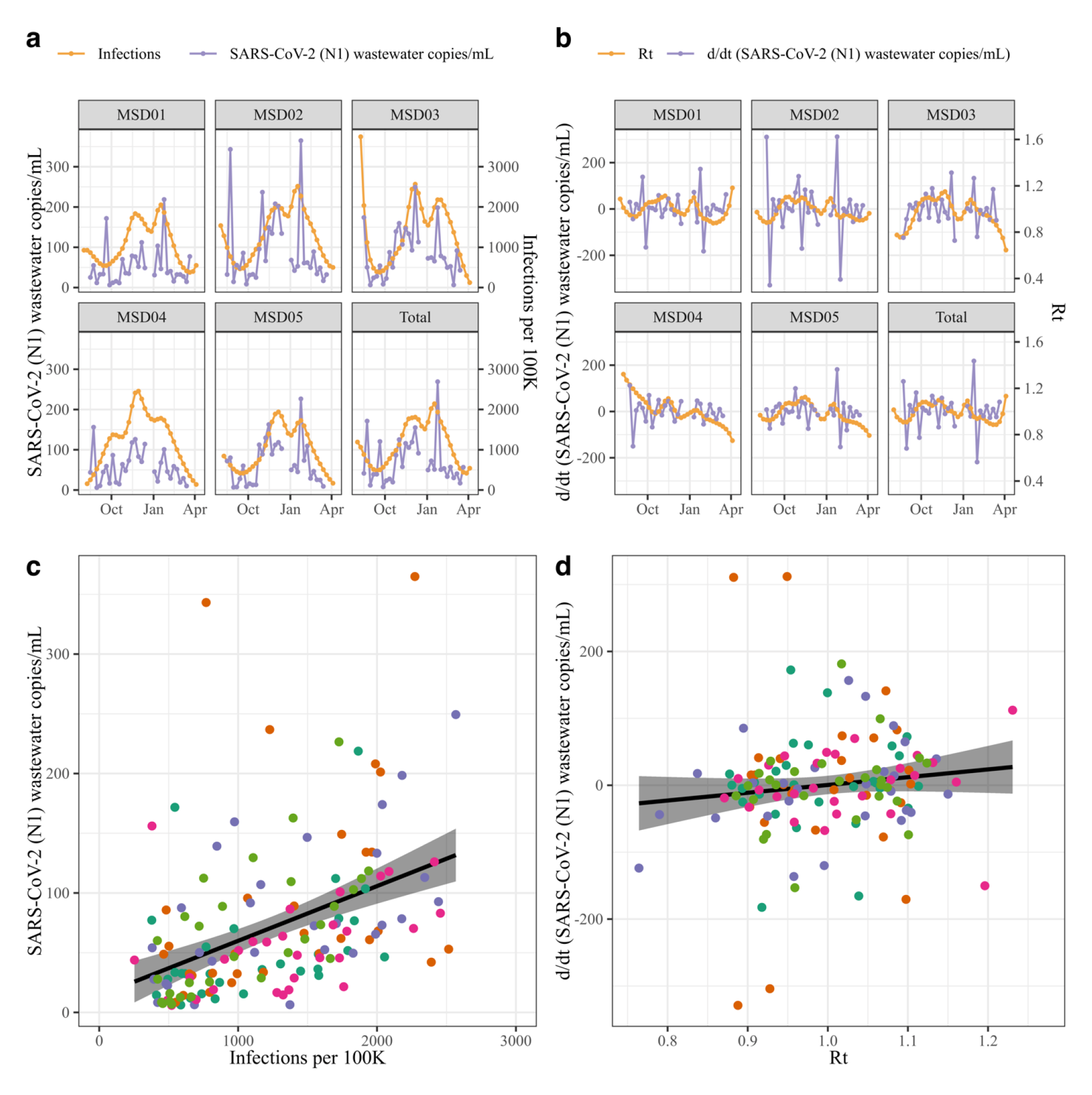
**Relationship between wastewater (N1) data and modeled estimates of infections and *Rt.*** (ab) Relationship between infections and SARS-CoV-2 N1 (copies/mL); (cd) Relationship between *Rt* versus SARS- CoV-2 N1(copies/mL, first order difference). (ac) Overlapping timeseries of wastewater and epidemic outcomes; (bd) Scatterplot and fitted linear model.

### Case reports

We used daily case reports from Louisville Metro Department of Public Health and Wellness (LMPHW), aggregated by week and geocoded to sewersheds based on reported address or zip code. The first reliable reports were available for the week starting on July 6, 2020. The last available weekly report was for the week starting on March 29, 2021 (ending April 4, 2021).

### Death reports

LMPHW provided publicly reported death data on August 17, 2022. Data were filtered for August 1, 2020 to March 31, 2021. Total deaths for this period were 1090. Nearly 90% (978/1090) of the death records had address information within one of the five sewersheds. For the 112 reports that could not be assigned to a sewershed area, due to either absence of address and/or zip code information or a recorded address outside the county treatment plant area, we probabilistically assigned these deaths to one of the five sewersheds, weighted by the relative population sizes. Deaths were aggregated to weekly rates, matching the frequency of the observed case data.

### Wastewater data

Wastewater samples were collected two to four times a week (Table S2), between August 2020 through March 2021 and analyzed for SARS-CoV-2 (N1 gene) and the fecal indicator pepper mild mottle virus (PMMoV) by RT-qPCR (copies/milliliter (mL)).^18^ Weekly averages were computed from quantifiable data. We used raw data (copies of N1/mL) for our analyses. Wastewater data for the last two weeks of December 2020 were not available due to a laboratory closure.

### Serosurvey data

A stratified simple random sampling serosurvey was executed in the study area over four discrete time periods.^19^ We used aggregated data, by wave and sewershed area at the four dates within the current study period. These data contain the number of serosurvey samples taken in a wave for each sewershed, the number of positive samples, and a weighted estimated percentage (with 95% CI) of the seropositivity in the population. We used these data to validate the results from our model analyses.

### Ethics

For the seroprevalence and data on COVID-19 deaths and infected individuals provided by the LMPHW under a Data Transfer Agreement, the University of Louisville Institutional Review Board approved this as Human Subjects Research (IRB number: 20.0393). For the wastewater data, the University of Louisville Institutional Review Board classified this as non-human subjects research (reference #: 717950).

## Model

### Analytic scenarios

We modeled four combinations of input data: (1) a ‘Cases-Deaths Model’, using cases and deaths data, consistent with the published version of the nowcasting model; (2), an ‘Additive Model’, using wastewater data in addition to cases and deaths data; (3) a ‘Substitutive Model’, using wastewater and deaths data; and (4) a ‘Deaths-Only Model’, using only deaths data, representing a worst-case scenario where no wastewater or case data are available. We fitted the four models to the timeseries data for each of the five sewersheds as well as their combined aggregate (Total), rendering six geographies. We ran the four models for each of the six geographies for each cumulative month of data after an initial first two months of data. There were 8·5 months of data available, and we created monthly snapshots of the data from month 2 to month 8, as well as the complete 8·5 months. We compared the Cases-Deaths Model to the Additive Model to assess the effects of adding wastewater data to existing case and death data. We compared the relative performance of the Substitutive Model and the Deaths-Only Model to the Cases-Deaths Model to assess the effects of including wastewater data when case data are absent. Finally, we compared the deviation of each of the five sewersheds from the Total to assess geographical variation.

### Nowcasting model

We used a published Bayesian mathematical back-calculation model that estimate SARS-CoV-2 infections and transmission rates from observed case and death data.^20^ We modified this model to work with weekly data, and to use wastewater data as an additional input (Supplementary Methods). To implement models that include wastewater (Additive and Substitutive Models), we first determined the transformation of wastewater data with the strongest correlation with the modeled estimates from the Cases-Deaths Model across the five sewersheds. We correlated the raw measurements of the N1 gene (copies per mL) with the infection estimates, and correlated the first-order differenced wastewater data (rate of change in the wastewater levels) with the estimates of *Rt*. The best fitting model informed our implementation of the model including wastewater.

## Outcomes

We compared predictive accuracy of the infections and *Rt* nowcasts between models, using four measures used by the COVID-19 Forecast Hub to evaluate the accuracy of forcasts (preprint),^21, 22^ adapted to compare two predictions rather than a prediction to a ground truth (Supplementary Methods): (1) the Absolute Difference between the point estimates (median of posterior distribution) of two models. A smaller Absolute Difference indicates a better agreement of two models; (2) the Sharpness, which is the weighted average of the widths of Credible intervals across K=11 coverages (10%, 20%, 30%, …, 90%, as well as 95% and 98%), with weights of 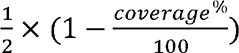. A smaller Sharpness indicates a more precise prediction; (3) the Coverage, defined here as the coverage of the 95% credible interval from one nowcasting model of the posterior distribution of the nowcasts from the comparison model; and (4) finally, the Weighted Interval Score (WIS), which is the weighted penalized average of the Absolute Difference of the K Credible intervals from the median estimate of the target model (Supplementary Methods).^21^ These measures were computed within models, that is, assessing the predictive value of each of the snapshot nowcasts to the estimates on that date from the complete data, and between models, that is, the difference between the two models, to quantify the agreement between models. We also assessed the historic reconstruction by comparing the historic estimates qualitatively by visual inspection, and quantitatively, by computing the overlap of the 95% credible intervals of the two posterior distributions. For models that included wastewater data (Substitutive and Additive models), we also assessed the fit of the modeled wastewater levels to the observed data.

### Sensitivity analysis

We tested the sensitivity of the results to outliers in the wastewater data, by refitting nowcasting models after removing outliers. To identify outliers, we fitted a smoothing spline to the daily wastewater timeseries data, and classified observations as outliers if their deviation from the spline was greater than three times the interquartile range of the deviations. Between 2% and 19% of the data points were marked as outliers (Table S2). We removed outliers, re-calculated weekly averages, and re-ran analyses on the adjusted data (Figure S1).

### Validation

To validate the nowcast estimates, we compared the cumulative infection estimates against the serosurvey data and against the estimates produced by the published nowcasting model using daily case and deaths data compiled by Johns Hopkins University, since the beginning of reporting up until December 2021.^23, 24^

We present results estimated for the Total sewershed, without outliers removed, and focus on the infections outcome. By default we present nowcast estimates for the complete timeseries, and refer to other snapshots where relevant. The results for individual sewersheds and each snapshot are available for both the infections and the *Rt* outcomes in the supplementary materials and referenced where relevant.

### Software

Data were analyzed using R (version 4.1.0) and the rstan package (version 2.21.5).^25, 26^ Figures were rendered using ggplot (version 3.3.6).^27^

## Results

### Correlation of wastewater with epidemiological outcomes

The timeseries of wastewater data correlated positively with the modeled infections (r = 0·362, [95% CI, 0·285 – 0·434], SARS-CoV-2 N1 outliers removed r = 0·486, [0·415 – 0·551], Figure 1ab). The first order difference of the SARS-CoV-2 N1 timeseries had a weak correlation with *Rt* estimates (r = 0·032 [-0·057 – 0·121], outliers SARS-CoV-2 N1 removed r = 0·092, [-0·003 – 0·185], Figure 1cd). Based on these results, we modeled the wastewater data assuming a linear relationship to the infection estimates (Supplementary Methods).

### Predictive performance of each model

In comparison to the Deaths-Only model, the addition of case data (Cases-Deaths model) resulted in smaller Sharpness, Absolute Deviation and WIS for within model predictive performance (Figure S2, Table 1). Similarly, a model fit to cases, deaths and wastewater data had improved performance metrics relative to a model using only deaths and wastewater data (Additive versus Substitutive Models). The addition of wastewater data resulted in improved performance metrics when case data were present (Additive Model versus Cases-Deaths Model), but not when case data were absent (Substitutive Model versus Deaths-Only Model). The relative reductions in Absolute Deviation, Sharpness and WIS were larger for the addition of case data than for the addition of wastewater data. The coverage across the snapshots of the complete data was similar for each of the four models.

**Table 1:** Predictive and historic performance of log(infections) nowcasts, within and between models.

| Comparison | Statistic | Cases-Deaths | Additive | Substitutive | Deaths-Only |
| --- | --- | --- | --- | --- | --- |
| Within model predictive performance | Absolute Deviation | 0.325 | 0.291 | 0.757 | 0.667 |
|  | Sharpness | 0.0168 | 0.0165 | 0.0435 | 0.0477 |
|  | Coverage | 99.3% | 99.3% | 99.3% | 99.3% |
|  | WIS | 0.223 | 0.208 | 0.517 | 0.500 |
| Between model predictive performance | Absolute Deviation | - | 0.246 | 1.18 | 1.35 |
|  | Sharpness | 0.0168 | 0.0165 | 0.0435 | 0.0477 |
|  | Coverage | - | 93.9% | 98.7% | 96.9% |
|  | WIS | 0.149 | 0.188 | 0.691 | 0.789 |
| Between model historic reconstruction | Absolute Deviation | - | 0.280 | 0.321 | 0.397 |
|  | Sharpness | 0.0069 | 0.0064 | 0.0112 | 0.0125 |
|  | Coverage | - | 76.8% | 91.2% | 92.5% |
|  | WIS | 0.059 | 0.156 | 0.185 | 0.231 |

For within-model comparisons, the last estimates from each model using the snapshots are compared against the timeseries estimates from the same model using the complete data. For the between model comparison of predictive performance, each the last estimates from each snapshot from each model are compared against the last estimates from the same snapshot of the Cases-Deaths Model. For the between model historic reconstruction, the timeseries estimates from each model using the complete data are compared against the estimates from the Cases-Deaths Model using the complete data.

### Addition of wastewater

Using wastewater data in the nowcasting model in addition to the cases and deaths data or to the deaths data did not result in any qualitative differences in the timeseries of infection estimates (Figure 3A). While the 95% credible intervals of the Additive Model’s historic reconstruction of infections covered only 76·8% of the Cases-Deaths Model’s posterior distribution, across the snapshots, the 95% Credible intervals of the last date’s nowcasts covered on average 93·9% of the Cases-Deaths Model’s posterior distribution. The Additive Model estimated a higher overall incidence of infections, but this difference was not statistically significant. Similarly, in the Substitutive Model the estimated incidence of infections was higher than in the Deaths-Only Model.

**Figure 3.**
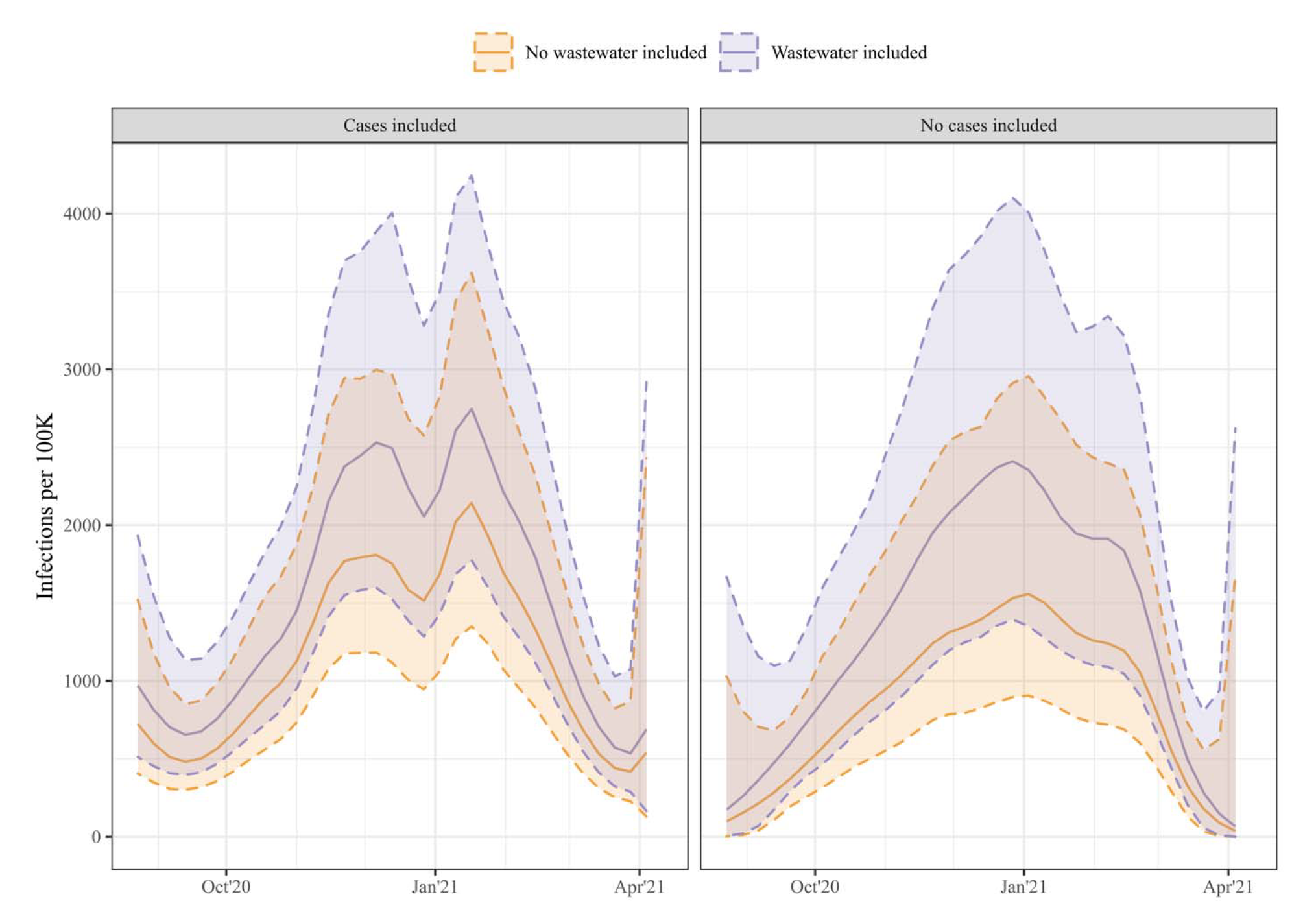
**Modeled infections per capita timeseries for the total sewershed.**

**Figure 4.**
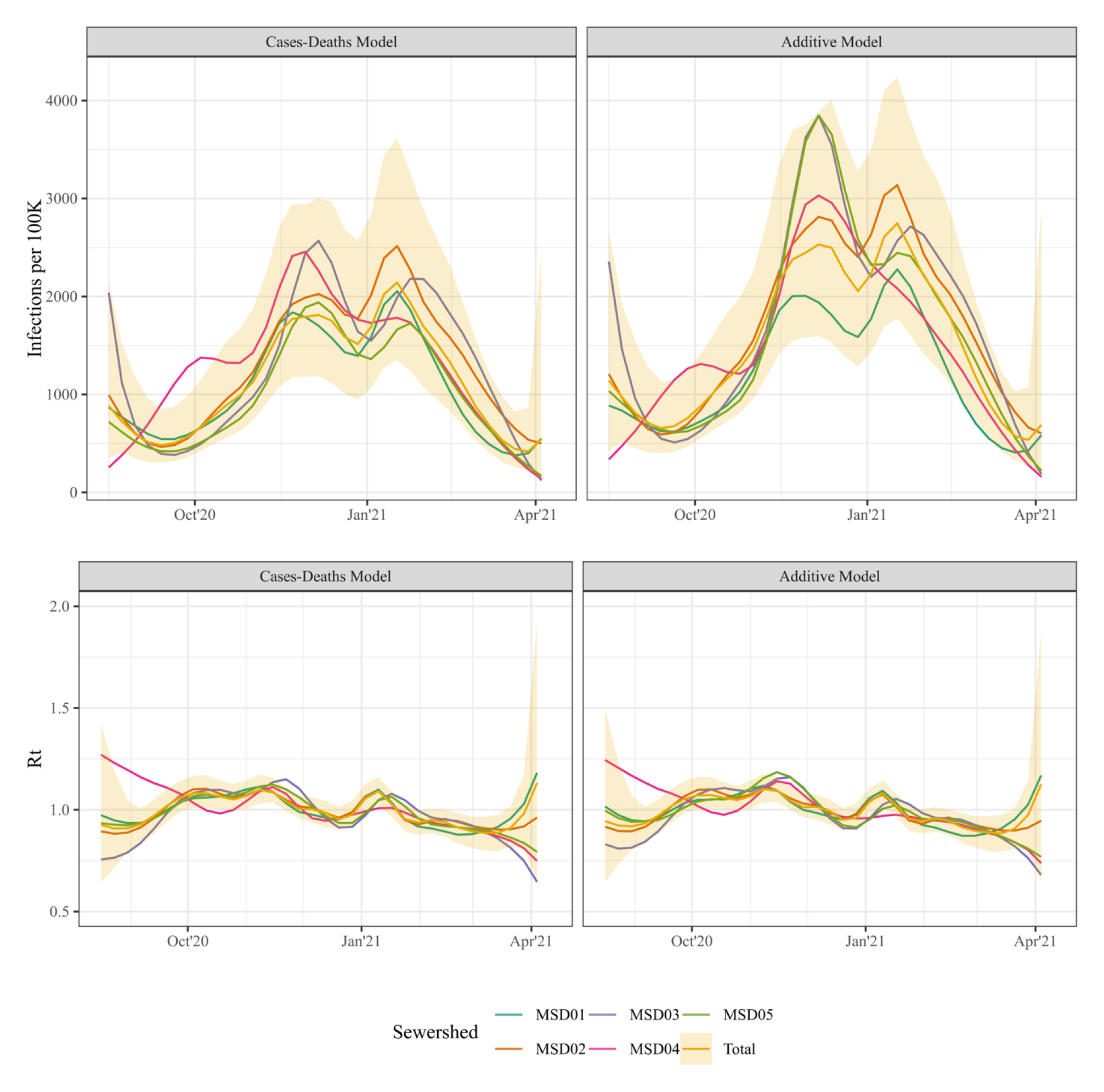
**Infection and *Rt* estimates for each sewershed.**

We found no strong differences in the within-model predictive performance of the Additive Model and the Cases-Deaths Model. The Additive Model had slightly lower scores on the Absolute Deviance, Sharpness and WIS, indicating stronger internal consistency of the estimates as data accrued. Out of the three alternatives to the Cases-Deaths Model, the Additive Model has the lowest WIS scores, indicating the best tradeoff in precision and certainty relative to the Cases-Deaths Model.

### Substitution of wastewater

The Deaths-Only Model outperformed the Substitutive Model in the within-model assessment. While the average Coverage of the historic last estimates to the complete estimates was similar, Absolute Difference, Sharpness and WIS were less for the Deaths-Only Model (Table 1), indicating a more consistent prediction when only deaths data were used compared to wastewater and deaths data. However, relative to the Cases-Deaths Model, the Substitutive Model had slightly better predictive and historic reconstruction power. The Absolute Difference of the median log(infections) from the Substitutive Model to the Cases-Deaths Model across historic runs was 0·321, while the Deaths-Only Model has an Absolute Deviation of 0·397 (Table 1). The WIS was lower for the Substitutive Model than the Deaths-Only Model both when comparing to the Cases-Deaths Model across the snapshots and in the historic reconstruction.

### Geographic granularity and wastewater

The 95% Credible Interval of the timeseries estimates for the complete county covered MSD02 best (Coverage of 89·8% for the Cases-Deaths Model, and 89·6% for the Additive model), and MSD04 worst (Coverage of 69·7% and 72·1% respectively; Table 2, Figure 3). While the historic reconstruction of the timeseries of infections and *Rt* was on average not statistically different, smaller population level sewersheds had larger Absolute Deviations and lower Coverage from the Total estimates. Notably, for MSD01, the largest sewershed in terms of population and area, the Coverage of the Total estimates was lower and the WIS was higher for the Additive Model compared to the Cases-Deaths Model, indicating that the wastewater data increased the variability in estimates between the sewersheds.

**Table 2.** Historic overlap of log(infections) estimates for each sewershed to the estimates for the total sewershed.

| **Model** | **Statistic** | **MSD01** | **MSD02** | **MSD03** | **MSD04** | **MSD05** |
| --- | --- | --- | --- | --- | --- | --- |
| Cases-Deaths Model | Absolute Deviation | 0.106 | 0.157 | 0.178 | 0.330 | 0.206 |
|  | Sharpness | 0.0092 | 0.0093 | 0.0113 | 0.0121 | 0.0142 |
|  | Coverage | 89.5% | 89.8% | 82.2% | 69.7% | 70.4% |
|  | WIS | 0.086 | 0.098 | 0.127 | 0.206 | 0.131 |
| Additive Model | Absolute Deviation | 0.270 | 0.136 | 0.185 | 0.289 | 0.213 |
|  | Sharpness | 0.0088 | 0.0085 | 0.0097 | 0.0108 | 0.0110 |
|  | Coverage | 77.1% | 89.6% | 80.9% | 72.1% | 75.5% |
|  | WIS | 0.156 | 0.089 | 0.127 | 0.185 | 0.131 |

All statistics are calculated comparing the historic estimates from the complete data for each sewershed to the historic estimates from the complete data for the total area. Coverage is defined as the average percentage of the posterior distribution from each sewershed covered by the 95% Credible Interval from the Total dataset.

### Validation

Modelled cumulative infection estimates were consistently higher than both the seroprevalence estimates derived from survey data (Figure 5), and the modelled cumulative infection estimates for the entire county (based on cases and death data since the beginning of the pandemic).^24^ For MSD02-05 the serosurvey data and the cumulative infection estimates follow a similar trend, while for MSD01 the serosurvey data flattens out over the last two observations, while the modelled estimates continue to increase.

**Figure 5.**
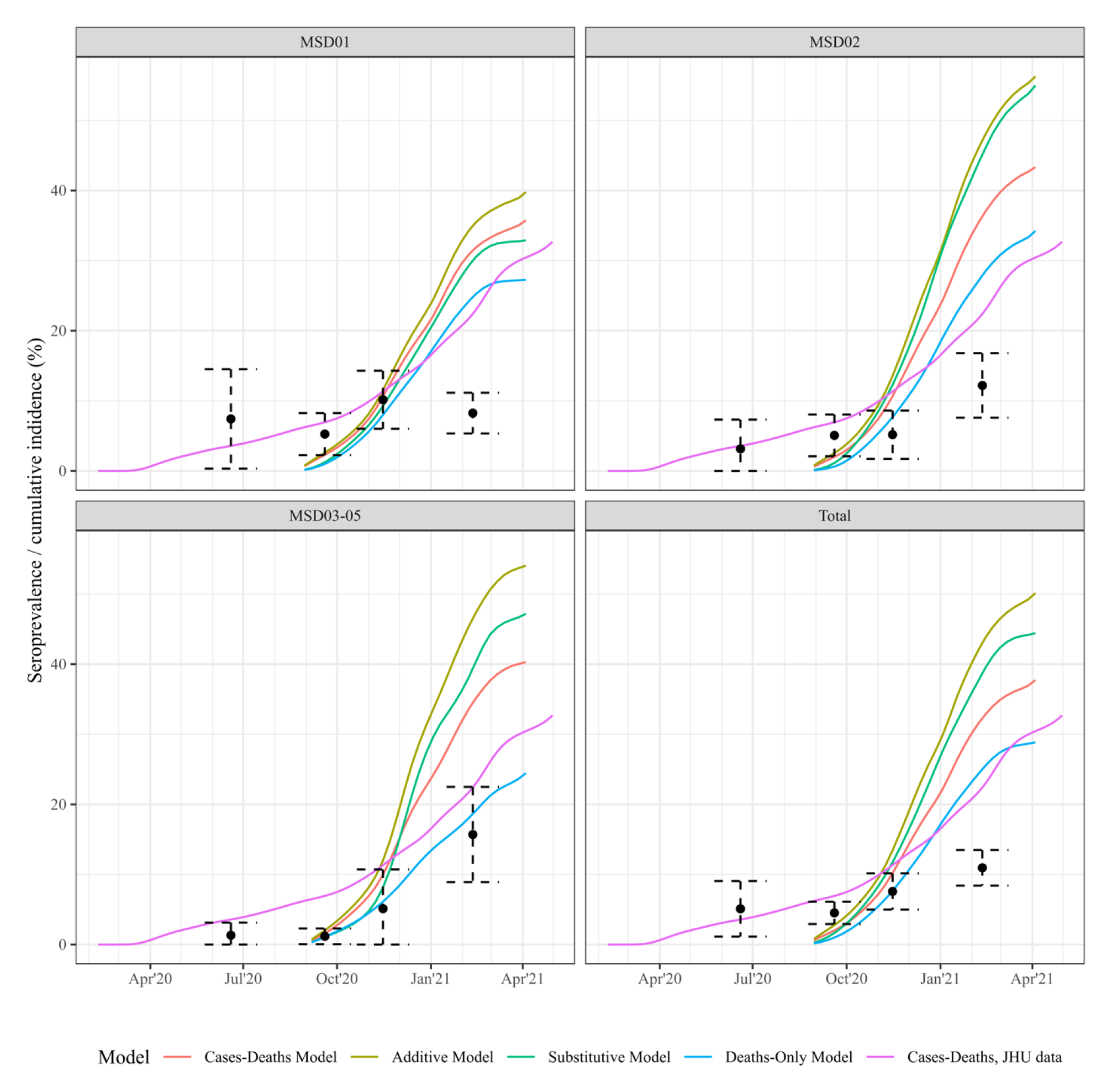
**Serosurvey and cumulative incidence estimates for Jefferson County, KY (USA).**

## Discussion

This study assessed the value of wastewater data for infectious disease nowcasting. We used SARS-CoV-2 case, death and wastewater data from Jefferson County, Kentucky and an adapted version of a published nowcasting model to evaluate the predictive and historic performance of wastewater data if they were used in addition or as a replacement for case data.

The results of our study showed a positive association between wastewater data and infection estimates from the nowcasting model fit to cases and death data. There were no significant differences in the within-model predictive performance, other than the expected improvements in Sharpness and WIS as more data were included. The addition of case data resulted in a greater relative improvement in the performance than the addition of wastewater data. The model containing all three data sources had the highest predictive performance. Nonetheless, the additive benefit of wastewater data was limited, as these data did not improve the predictive performance substantially.

This work demonstrates that wastewater data may be a viable substitute for case data, as estimates using death and wastewater data approximated the estimates from a cases and deaths model closer than when only deaths were used. As the availability of traditional surveillance data deteriorates during an ongoing pandemic, the possibility of using wastewater data in nowcasts and monitoring transmission is of high publich health interest and underlines the potential utility of these data for future pandemic preparedness. This approach will have additional relevance in low-resource settings where systematic collection of epidemiologic data is likely to be severely limited. Such low-resource areas are likely not only low-and middle-income countries, but rural areas within both the United States and Europe. Further research is needed to examine the relationship between wastewater and infections across multiple locations and longer timeseries to confirm our findings and to evaluate their applicability in different social and geographic contexts.

We found that the models that include wastewater data consistently estimated higher infections than the models without wastewater data included, which may be an artefact of a different estimated probability of progressing to symptomatic and severe disease. Additionally, we considered the question of the potential use of wastewater in smaller populations (sewersheds), where count surveillance data (e.g., case, death, hospitalizations) may not be as readily available as in larger countywide areas, yet wastewater data can still be obtained. The epidemiological trends estimated for the smaller sewersheds in our study area deviated more from the aggregate estimates than the larger sewersheds. This highlights the importance of high frequency local surveillance data for guiding future public health responses, as transmission may differ by area, and local granular data might help monitor disease trends more closely.

This study is a first attempt to assess the value of adding wastewater data in a nowcasting model. Several limitations should be considered. First, the study had a limited scope in time and geography, and we only considered a single nowcasting model. When we compared estimates of cumulative infection generated by this study to other estimates, we found sewershed-specific estimates as well as the aggregated estimates exceeded the estimates from the nowcasting model using a longer case and death timeseries, indicating the contribution of historical transmission to cumulative infection estimates. The sewershed specific estimates consistently exceeded the measured seroprevalence data; seroprevalence data only included the population older than 18 years whereas wastewater captures a wider portion of the population. The nowcasting model did not account for multiple infections. At the end of the period under consideration, the estimated cumulative infections were around 2-3 times higher than the estimates from the seroprevalence survey, consistent with earlier findings.^19^ It is unclear whether this reflects overestimation on the part of the model, or losses in seropositivity among previously infected individuals.

Another potential set of limitations is linked to our decisions regarding outliers and not normalizing the wastewater data. Past WBE research demonstrates a wide range of quantification and normalization methods, and normalization may not improve the signal in the wastewater data.^28, 29^ Across the sewersheds, up to 19% of the wastewater observations could be marked as outliers using a spline timeseries. Wastewater data may detect local events, such as festivals or conferences, which may be an indication of transmission at a short timescale, but not of infections or sustained transmission in the population. This could have been a factor in the current data, as the Kentucky Derby took place in Louisville, Kentucky on September 5, 2020, which coincides with some of the extreme datapoints in the wastewater data. Nonetheless, the sensitivity analyses that excluded wastewater data outliers did not result in different conclusions in this study.

One potential reason for the limited additive value of the wastewater timeseries is that these data were much less smooth compared to the case and death data (Figure S1). In other words, while adding information, the data also introduced further uncertainty. For future use of wastewater data in public health surveillance, it is important to investigate the sampling frequency and data quality needed.

The results from this study of a strong correlation between the wastewater data and estimated infections correspond to other studies where wastewater data were used to predict case or hospitalization reports.^12, 14^ The strength of this relationship appears to be stronger when daily moving averages of the wastewater data are available. ^25^ Despite potential bias introduced by using the wastewater data to inform the parameterization of those data in the model, the additive and substitutive power in this study were not very strong, when compared with a preprint study in nowcasting SARS-CoV-2 infections in the Boston metropolitan area, covering a much larger population.^13^ This further supports the need for future research into the conditions required for using wastewater data in infectious disease nowcasting. Finally, our models included a simple linear relationship between wastewater data and modeled infections. There is a range of additional assumptions and more complex modeling choices that can be made to further support the use of wastewater in nowcasts.^30^ This might complicate models and might make them less versatile across various infectious diseases, as these assumptions are disease specific, or require much additional data, like temperature and flow rates. The presence and level of viral load in wastewater data is a function of many other variables (such as temperature, sample type, flow rates, distance from households), and the association between the viral load and the transmission in the population is a function of the amount and length of viral shedding at various disease stages (preprint).^31^

In conclusion, in this case study, we found that the use of wastewater data improved the performance of COVID-19 nowcasts for Jefferson County, Kentucky. However, these improvements were modest, particularly when case data were available. Future research on the value of wastewater data for nowcasting when case data are absent or unreliable would be beneficial. For public health officials this is critical information in balancing the focus of surveillance efforts – while wastewater may offer early detection, it’s incremental value for nowcasting may be limited if high-quality case reports are available, providing a reminder of the value of investments in traditional surveillance data. It is also possible that wastewater data will provide stronger evidence in other settings, and research on approaches to strengthen the value of these data for surveillance proposes is important for ongoing pandemic preparedness.

## Data Availability

All data produced in the present study are available upon reasonable request to the authors

