## Supplement for "Predictive power of wastewater for nowcasting infectious disease transmission: a retrospective case study of five sewershed areas in Louisville, Kentucky"

**Table of Contents**

Supplementary Methods ------ page 2-3
Supplementary Tables
Table S1 --------------page 4
Table S2 --------------page 5
Supplementary Figures
Figure S1 -------------page 6
Figure S2a-f ---------pages 7-12
Figure S3a-f ---------pages 13-18

### Supplementary Methods

#### Adjustments to covidestim model

We used the model described in Chitwood et al. (2022) as the basis for the analyses, with a few adjustments made. First, we adjusted the timeframe from daily to weekly, to match the available data. We updated the prior distributions on the progression, diagnosis and reporting delays accordingly. Furthermore, we included a fraction of the population that was already infected on August 1, 2020, to account for the fact that not the entire population was available for infections. We used the fraction of the population infected in Jefferson County, as estimated by covidestim on August 1.

#### Model options

We considered two versions of including wastewater in the model, depending on the best fit to either the Rt or the infection estimates.

*Option 1: Rt and differenced wastewater data*

For this option, we considered that the differenced wastewater data might be informative of Rt. That is, the detectable changes in the wastewater data would be indicative of the changes in the force of transmission in the population. This option requires that the differenced wastewater is included in the model, as well as indicators of observed and non-observed data points. For the observed datapoints, a linear model with normal residuals would describe the relationship between modeled Rt and the observed differenced wastewater data. In other words, force of infection is a predictor of the observed change in wastewater data.

*Option 2: Infections and wastewater data*

For this option, we considered that the wastewater data might be linked to infections. That is, there is a linear relationship between the new infections in a week and the observed level of wastewater data. Specifically, we modeled this by a Student's T distribution with 10 degrees of freedom, to allow for potential outliers.

#### Equations for measures of performance evaluation

For the evaluation of the nowcast performance, we used four measures used in the CDC forecasting evaluation of COVID-19. We made slight adaptations to these calcualtion, as they are defined with regard to a ground truth, while we compared the predictions of two models. The calculations are described in the following section. For the sake of clarity, the equations are written for the comparison of Model X to Model Y, where Model Y is the 'target' model.

*Absolute Difference*

The Absolute Difference is the absolute difference between the posterior median of Model X, $\mu_x$ and the posterior median of Model Y, $\mu_y$:

$$Absolute\ Difference = |\mu_x - \mu_y|$$

*Sharpness*

The Sharpness is defined as the average width of the the credible interval of Model X. We consider K = 11 nominal credible intervals, $CI_k = \{10\%, 20\%, \dots, 90\%, 95\%, 98\%\}$, that are defined by lower and upper bounds $l = \frac{a_k}{2}$ and $u = 1 - \frac{a_k}{2}$, the quantiles of the posterior samples, where $a_k = \{.9, .8, \dots, .1, .05, .02\}$.

$$Sharpness = \frac{1}{K}\sum_{k=1}^{K}\frac{\left(u_x^{\{CI_k\}} - l_x^{\{CI_k\}}\right)}{\mu_x} \times \frac{a_k}{2}$$

*Coverage*

The Coverage of the 95% Credible Interval of Model X of the posterior samples from Model Y is defined as:

$$100\% \times \frac{1}{N}\sum_{n=1}^{N}\theta_y^n \times \mathbf{1}(l_x^{CI} < \theta_y^n < u_x^{CI})$$

Where $\mathbf{1}(l_x^{CI} < \theta_y^n < u_x^{CI})$is and indicator function, that is 1 if the posterior sample n from Model Y falls within the lower and upper bounds of the 95% Credible Interval from Model X, and 0 if not. Coverage can also be calculated as the percentage of posterior sampler from Model X that are covered by the 95% Credible interval from Model Y.

*Weighted Interval Score*

The Weighted Interval Score (WIS) is defined as the weighted average distance of each of the K Credible intervals from Model X from $\mu_y$, the median estimate of Model Y. It is calculated in two steps. First, the Interval Score (IS) is calculated:

$$IS_{a_k} = \left(u_y^{\{CI_k\}} - l_y^{\{CI_k\}}\right) + \frac{2}{a_k} \times \left(l_x^{\{CI_k\}} - \mu_y\right) \times 1\left(\mu_y < l_x^{\{CI_k\}}\right) + \frac{2}{a_k} \times \left(\mu_y - u_x^{\{CI_k\}}\right) \times 1(\mu_y > u_x^{\{CI_k\}}$$

Which is a sum of the width of the kth interval, plus the absolute over- or undershoot if $\mu_y$ falls outside of that interval, penalized by the inverse of the width of that interval.

The WIS then calculates a weighted average of these ISs and the Absolute Deviation:

$$\frac{1}{K + \frac{1}{2}} \times (w_0 \times |\mu_y - \mu_x| + \sum_{k=1}^{K} w_k \times IS_{a_k})$$

Where $w_0 = \frac{1}{2}$ and $w_k = \frac{a_k}{2}$

Table S1. **Characteristics of studied wastewater treatment sewersheds of Jefferson County, KY (USA).**

| **Site name** | **Population**[a] | **Area ($km^2$)** |
|---|---|---|
| MSD01<br>Morris Forman Water Quality Treatment Center (MFWQTC) | 349,850 | 280 |
| MSD02<br>Derek R. Guthrie Water Quality Treatment Center (DRGWQTC) | 295,910 | 332 |
| MSD03<br>Cedar Creek Water Quality Treatment Center (CCWQTC) | 55,928 | 80 |
| MSD04<br>Floyds Fork Water Quality Treatment Center (FFWQTC) | 32,460 | 88 |
| MSD05<br>Hite Creek Water Quality Treatment Center (HCWQTC) | 31,269 | 67 |

[a] Based on 2018 U.S Census Bureau American Community Survey (ACS). Income is mean median household.

Table S2**. Descriptives of outlier detection and removal.**

| **Site name** | **N observations** | **N outliers (%)** | **N weeks NA due to outlier exclusion (%)** |
|---|---|---|---|
| MSD01 | 79 | 10 (13%) | 3 (3%) |
| MSD02 | 47 | 9 (19%) | 3 (10%) |
| MSD03 | 47 | 1 (2%) | 0 (0%) |
| MSD04 | 47 | 3 (6%) | 1 (3%) |
| MSD05 | 47 | 6 (13%) | 2 (7%) |

N=number of; NA = Not available.

Figure S1. **Timeseries of weekly SARS-CoV-2 N1 data with/without outliers removed**

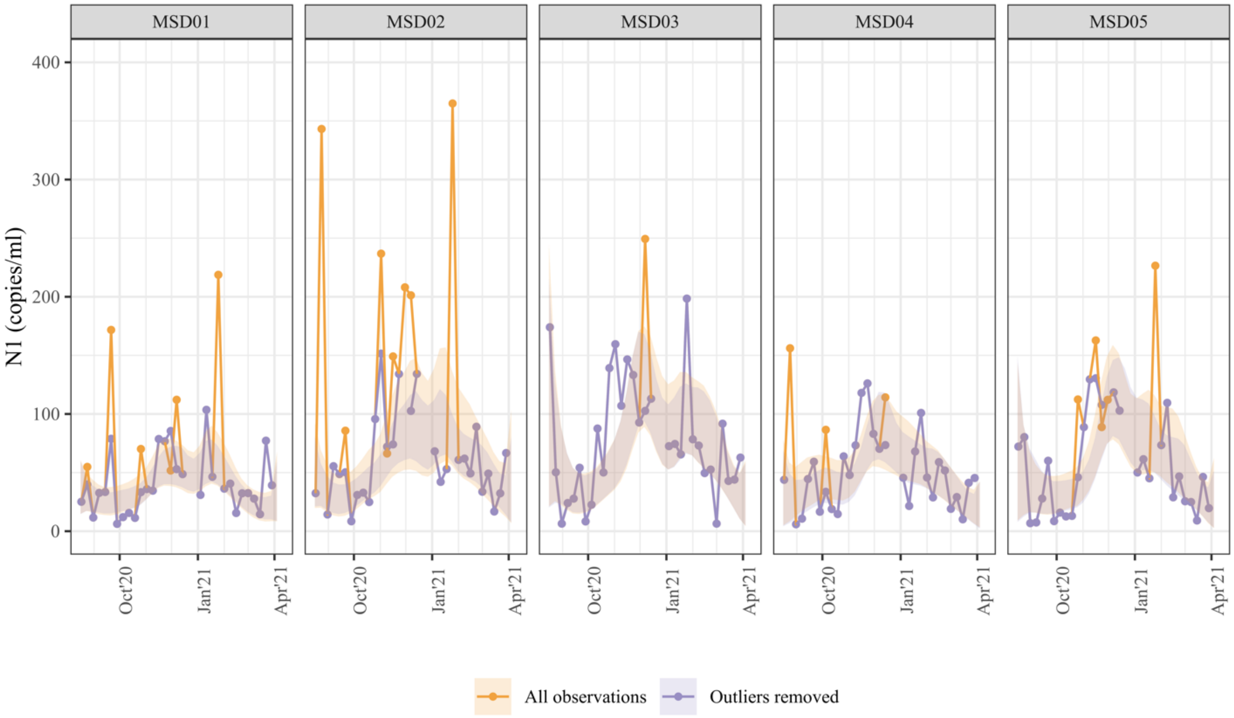

Figure S2. Sequential timeseries estimates of log(infections) estimates by model and sewershed.

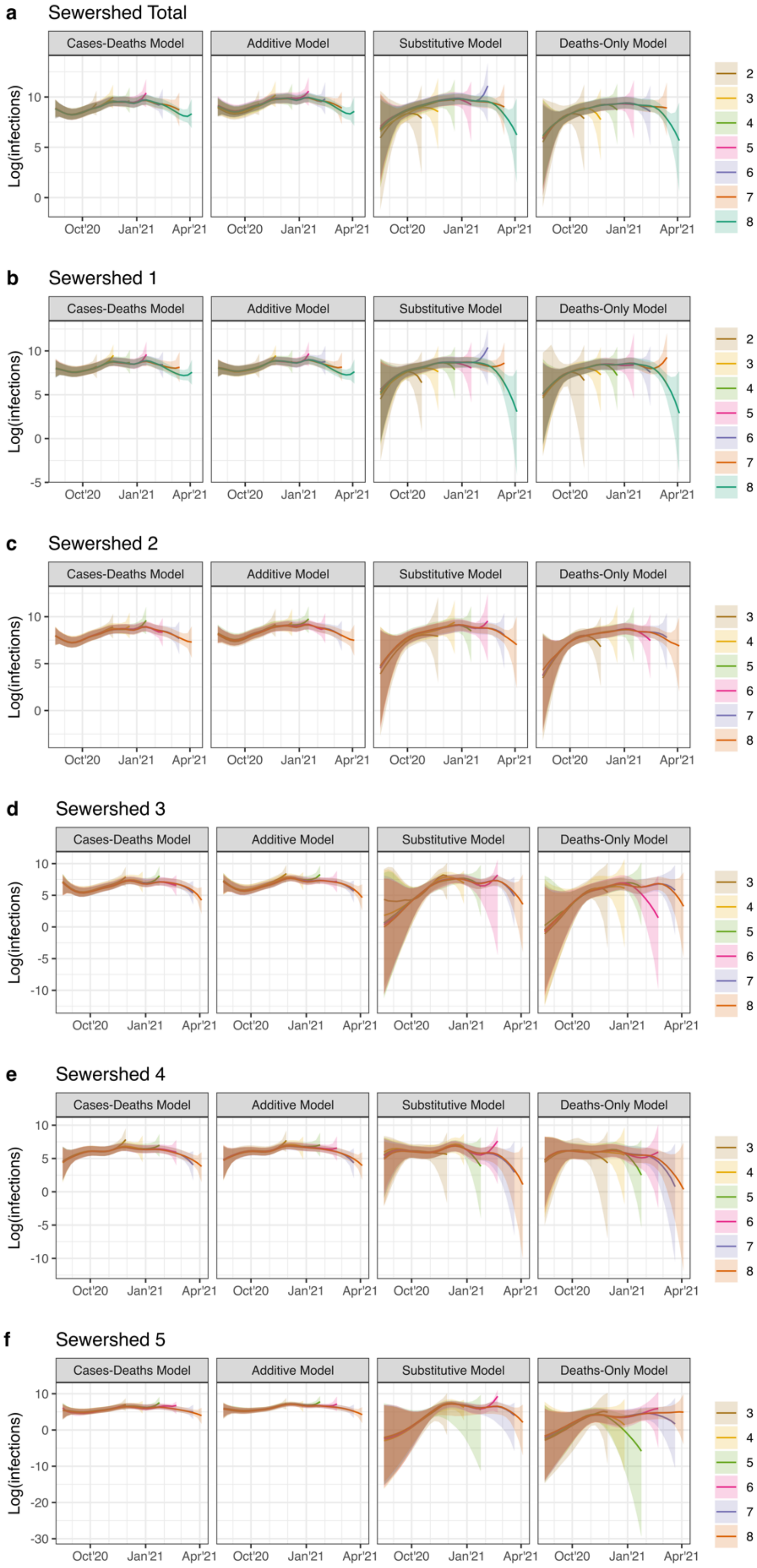

Figure S3. Sequential timeseries estimates of log($Rt$) estimates by model and sewershed.

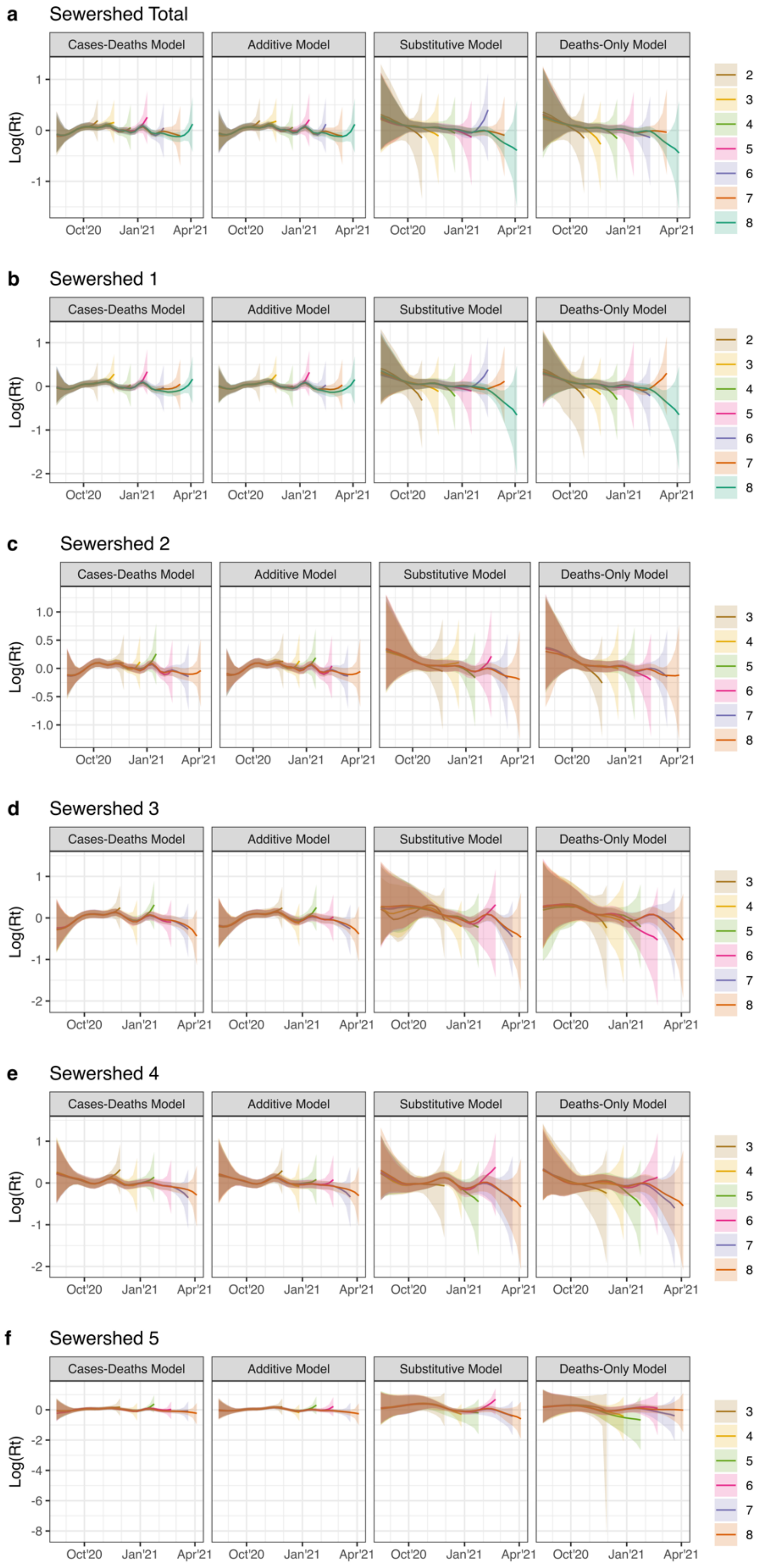
